# Speech, language, p-tau217 and neurofilament light chain in an Alzheimer’s Disease at risk population

**DOI:** 10.64898/2025.12.04.25341021

**Authors:** Alveena Siddiqui, Peter J. Snyder, Jessica Alber, Thayabaran Kathiresan, Adam P. Vogel

**Affiliations:** School of Health Sciences, The University Melbourne, Melbourne, VIC 3010, Australia; Redenlab Inc., Melbourne, VIC 3000 Australia; College of Pharmacy, The University of Rhode Island, Kingston, RI 02881, USA; Department of Neurology, Alpert Medical School of Brown University, Providence, RI 02903, USA; Memory and Aging Program, Butler Hospital, Providence, RI 02906, USA

**Author notes:** **Corresponding author**: Adam Vogel PhD, 550 Swanston Street, Parkville, VIC Australia 3010. A.P.V & P.J.S. are joint senior authors.

**Keywords:** Alzheimer’s disease, Speech and language biomarkers, p-tau217, Neurofilament light chain (NfL), Mild cognitive impairment (MCI)

## Abstract

**Introduction:** Alzheimer’s disease (AD) affects speech, language, and executive functions. Combining blood biomarkers with non-invasive speech analysis may aid early detection.

**Methods:** A speech battery was administered to three groups aged 65-70: cognitively normal low-risk (LRC), high-risk (HRC), and mild cognitive impairment (MCI).

**Results:** Variance in Mel-Frequency Cepstral Coefficients (MFCCs) predicted p-tau217 (F = 3.71, p = 0.005), with group-specific effects in LRC and HRC. In MCI, articulation rate, speech percentage, and pause percentage predicted p-tau217 (all p < 0.01). Language features including idea density, content density, and open-class word rate also predicted p-tau217 (all p < 0.05). MFCCs and syllable timing correlated with **NfL** (all p < 0.01). Lexical diversity differed between groups, notably in HRC and MCI.

**Discussion:** Speech timing, fluency, and voice quality, along with lexical richness and idea density, predicted p-tau217 and NfL, supporting acoustic and linguistic features as non-invasive digital biomarkers for early AD-related neurodegeneration.

## 1. Introduction

Alzheimer’s disease (AD) is a progressive neurodegenerative disorder marked by early disruption of memory, executive function, and language behavior ^1^. While current diagnostic frameworks increasingly incorporate fluid and imaging biomarkers of amyloid and tau pathology, there remains a pressing need for scalable, low-burden tools that capture clinically meaningful change across the disease spectrum. Speech and language analyses, non-invasive, repeatable, and sensitive to neurocognitive dysfunction, represent a promising set of digital tools for detecting early pathological changes and tracking disease progression ^2^.

Earlier work has reported that alterations in speech motor control (e.g., slowed articulation, increased pausing) and linguistic features (e.g., reduced idea density, lexical diversity) can distinguish individuals with MCI or early AD from healthy controls ^3–5^. These impairments reflect disruptions in temporoparietal, frontal, and subcortical networks vulnerable to AD pathology ^6^. However, few studies have directly linked speech and language features with established biomarkers of neurodegeneration and tau pathology in preclinical or prodromal stages ^2^.

Plasma biomarkers such as phosphorylated tau at threonine-217 (p-tau217) and neurofilament light chain (NfL) are increasingly adopted in both research and clinical contexts due to their accessibility and strong correlation with underlying neuropathology ^7^. P-tau217 closely reflects tau tangle accumulation and shows excellent discrimination of AD from other dementias ^8^, whereas NfL is a general marker of axonal injury and neurodegeneration ^9^. Elevated levels of NfL have been observed in AD and are associated with poor cognitive performance ^10^.

Evaluating their relationship with speech-derived metrics may illuminate the pathophysiological relevance of digital language markers and help refine endpoint selection in early-stage trials.

In this study, we examined the associations between digital speech and language features and plasma concentrations of p-tau217 and NfL across three participant groups: Low-Risk Cognitively normal (LRC), High-Risk Cognitively normal (HRC), and Mild Cognitive Impairment (MCI). Participants completed a structured battery of speech tasks, including sustained phonation, diadochokinetic repetition, reading, spontaneous monologue, and picture description. Acoustic and linguistic features were extracted using automated pipelines and evaluated using group comparisons and linear regression models.

We hypothesized that select speech and language features particularly those reflecting motor timing, lexical diversity, and informativeness would be significantly associated with biomarker levels, with distinct profiles observed across clinical risk groups. By establishing biomarker correlates of naturalistic speech, this work aims to support the integration of digital voice analytics into early detection and monitoring frameworks for Alzheimer’s disease.

## 2. Methods

### 2.1 Study Design and Participants

This was a cross-sectional analysis of speech and language features in relation to plasma biomarkers of Alzheimer’s disease pathology. Eligible participants were aged 55–80 years, fluent in English, and classified into one of three groups based on clinical and biomarker profiles. During the screening visit a cheek swab was obtained to determine apolipoprotein (APOE) genotype for assigning each subject to an appropriate group: Low-Risk Cognitively Normal (LRC), High-Risk Cognitively Normal (HRC), and Mild Cognitive Impairment (MCI). The LRC group included individuals without subjective cognitive complaints, no family history of Alzheimer’s disease, and did not carry an APOE ε4 allele. The HRC group comprised cognitively normal participants with subjective memory concerns, a positive family history, and they were all either hetero- or homozygous for the APOE ε4 allele. The MCI group included individuals with a clinical diagnosis of MCI, a Delayed Memory Index score ≤ 85 on the RBANS, and a positive Alzheimer’s biomarker result (either CSF or PET) showing abnormal beta-amyloid protein aggregation.

Exclusion criteria included history of neurological or psychiatric illness, current substance misuse, or uncorrected hearing or vision impairments.

### 2.2 Procedures

Participants completed a standardized speech battery, neuropsychological assessment (MoCA ^11^ and RBANS ^12^), and provided blood samples for analysis of plasma p-tau217 and neurofilament light (NfL) concentrations. Fasting blood samples (17.5ml blood including 10 ml for plasma and 7.5 ml for serum) were collected from every participant at baseline visit (V2). Plasma Ethylenediaminetetraacetic acid (EDTA) tubes were gently inverted 5-10 times and then centrifuged with a horizontal rotor for 10 minutes at 2000 x g within one hour of collection. After centrifugation, 1.0 mL aliquots of plasma were transferred to polypropylene tubes and stored at -80□C freezer within 2 hours of collection. Blood samples were sent to a central biorepository for analysis using the ultrasensitive immunoassay platform called the single molecular array (SIMOA platform) technology by Quanterix. Blood samples were analyzed for plasma p-tau217 (ALZpath p-tau217 assay), and NfL and then sent for measurement of p-tau217 and NfL level in plasma.

### 2.3 Speech and Language Tasks

Speech recordings were recorded using a lavaliere microphone coupled with an iPad using the Redenlab ® Mobile or Online applications at a sampling rate of 44.1 kHz. All recordings were conducted in a quiet, acoustically controlled room. Participants completed six standardized tasks that fit along a continuum of cognitive-motor complexity ^13,14^:

- **Sustained vowel phonation (AAAH)**: sustained /a:/ for at least 5 seconds in a clear voice
- **Diadochokinetic task (PTKA)**: rapid alternating production of the syllables /p/, /t/, /k/ for 10 seconds
- **Reading of a standard passage**: phonetically balanced Grandfather Passage ^15^
- **Monologue**: open-ended response describing a recent personal event
- **Days of the week recitation**: beginning with Monday
- **Picture description**: response to a picture description tasks where participant described what they could see in a fishing scene (see Figure S1 in Supplementary Materials)

The stability of measures derived from these tasks has been explored previously ^16,17^.

Acoustic signal processing was conducted using Redenlab Analyze (v0.14.42). Features extracted included harmonics-to-noise ratio (HNR), Mel-frequency cepstral coefficients (MFCC1–MFCC4), articulation rate (ARATE), speech time percentage (SPEECH_PERCENT), pause percentage (P_PERCENT), syllable count (SYL_COUNT) and duration metrics, and variability measures (e.g., coefficient of variation for pitch, energy, and syllables). Feature extraction followed previously validated procedures and included automated quality checks to exclude artifacts (Table 1)

**Table 1:**
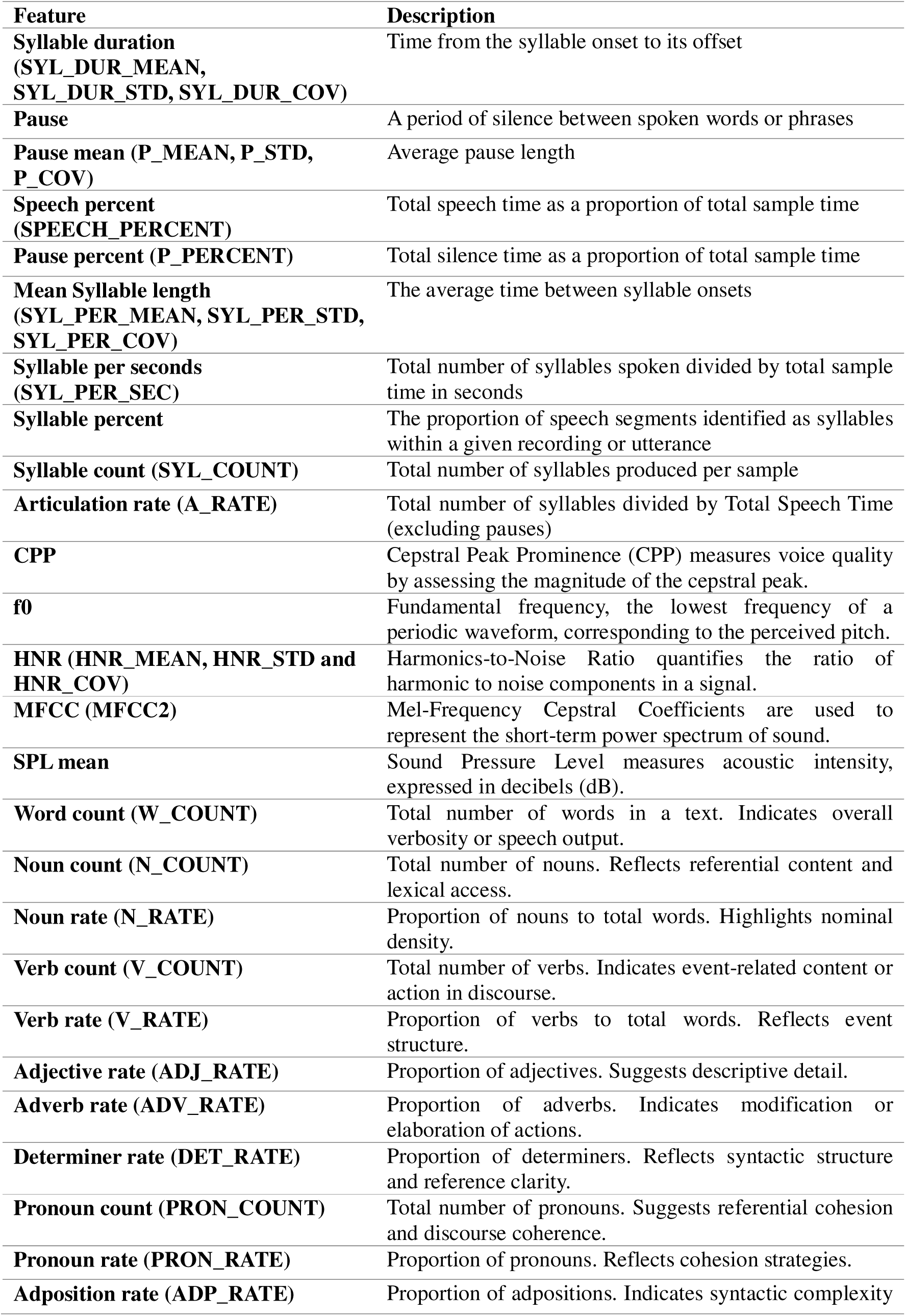
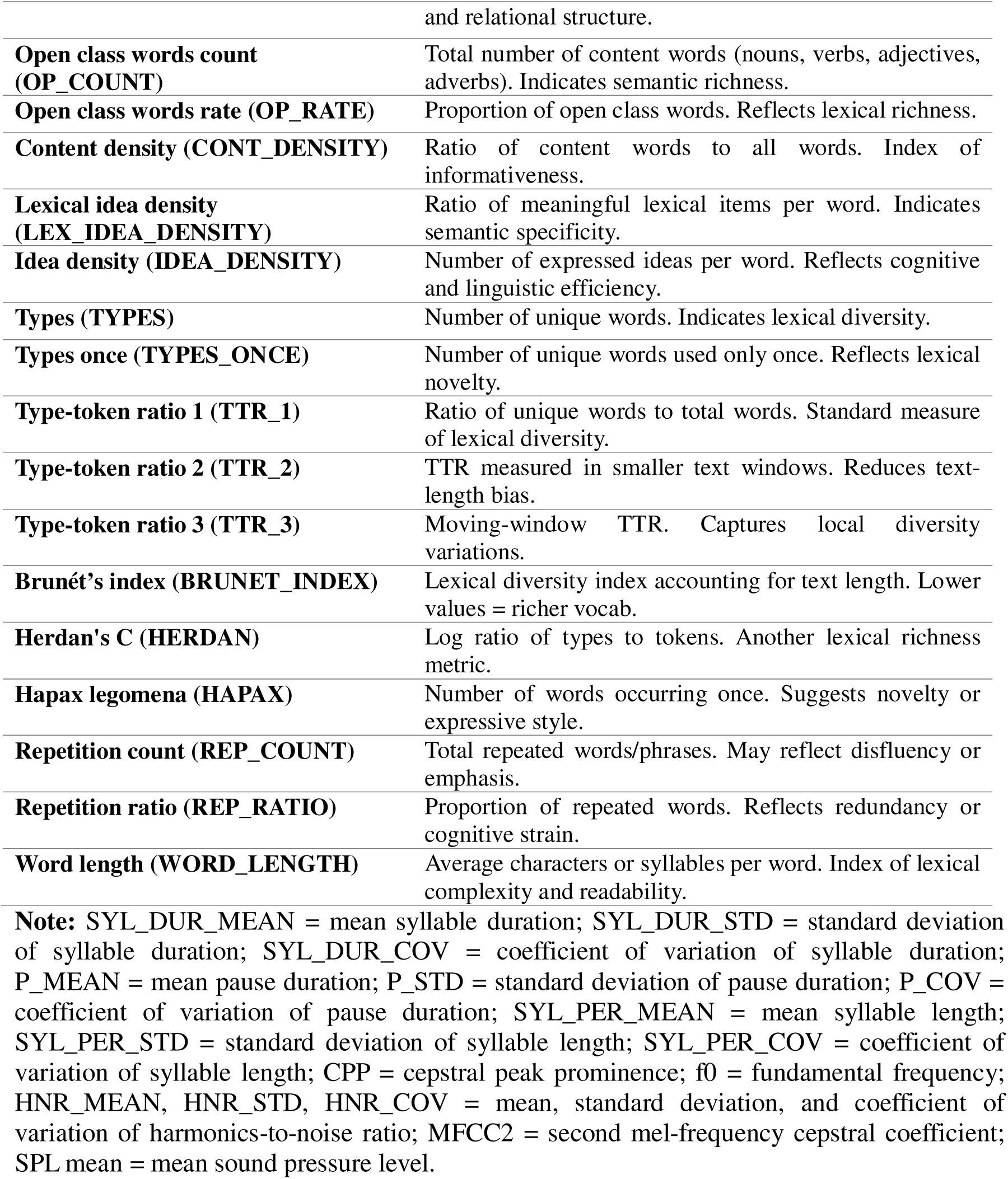
Motor speech, voice and language features extracted for analysis.

Linguistic analysis was performed on transcribed samples from the monologue and picture description tasks. Transcriptions were segmented and processed through a custom natural language processing pipeline. Extracted features included:

- **Syntactic and lexical diversity**: HERDAN, HAPAX, number of types and tokens
- **Grammatical composition**: noun count (N_COUNT), pronoun count (PRON_COUNT), adjective rate (ADJ_RATE), open/closed class ratios
- **Information metrics**: idea density (IDEA_DENSITY), lexical idea density (LEX_IDEA_DENSITY), content density (CONT_DENSITY), overt pronoun rate (OP_RATE)

Linguistic features were calculated using Stanza (v 1.10.1) and custom scripts written in Python (v 3.13.3), incorporating lemmatization and part-of-speech tagging aligned with Universal POS tags (Table 1).

### 2.4 Statistical Analysis

A series of linear regression analyses were performed to evaluate the relationship between speech and language features, and biomarkers (ptau-217 and NfL) in individuals categorized by group (LRC: low risk cognitively normal, HRC: High risk cognitively normal and MCI: Mild cognitively impaired). Analysis was performed using RStudio’s version 4.4.1 (2024-06-14) tidyverse, broom and gt libraries. Visualizations were created for statistically significant results using geom_smooth (method = “lm”) from the ggplot2 package. All data preprocessing and visualizations were conducted in R (version 4.4.1; R Core Team, 2024) using RStudio (version 4.4.1)

A complete-case approach was applied to exclude missing values. Separate linear regression models were constructed for each independent variable (speech and language features mentioned in methodology section) with interaction terms for “Assigned Group” to investigate group-wise differences. The dependent variables were ptau-217 and NfL. Model summaries were generated to report coefficients, 95% confidence intervals, and p values. Analytic measures metrics included F-statistics and adjusted R^2^. A threshold of p <0.01 was chosen to limit Type 1 error based on a higher number of variables, multiple interactions, and the risk of false positives.

Descriptive statistics summarizing participants’ demographic and clinical characteristics are provided in Table 2. Speech and language results are detailed in Table 3 (for p-tau217) and Table 4 (for NfL), with scatterplots illustrating significant regressions presented in Figures 1 and Figure 2. Supplementary Table S1*(a)* to S1*(d)* and S2*(a)* to S2*(l)* provide full results.

**Figure 1:**
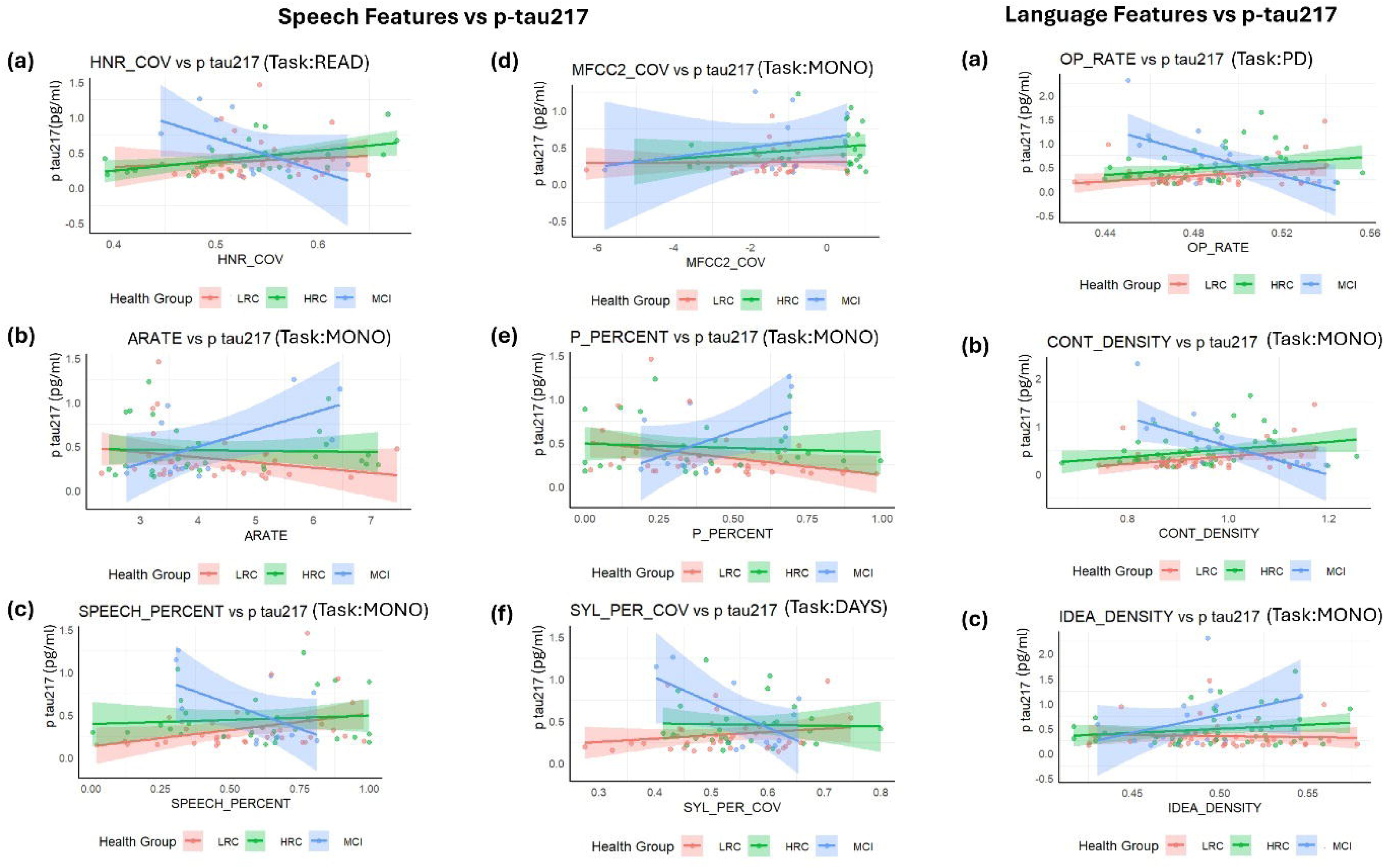
Regression plots for speech and language features with ptau-217. HNR_COV, harmonics-to-noise ratio coefficient of variation; MFCC2_COV, second mel-frequency cepstral coefficient coefficient of variation; OP_RATE, open class word rate; ARATE, articulation rate; P_PERCENT, pause percentage; CONT_DENSITY, content density; SPEECH_PERCENT, percentage of speech; SYL_PER_COV, syllable onset time coefficient of variation; IDEA_DENSITY, idea density. MONO, monologue; PD, picture description .LRC, low-risk cognitively normal group; HRC, high-risk cognitively normal group; MCI, mild cognitive impairment.

**Figure 2:**
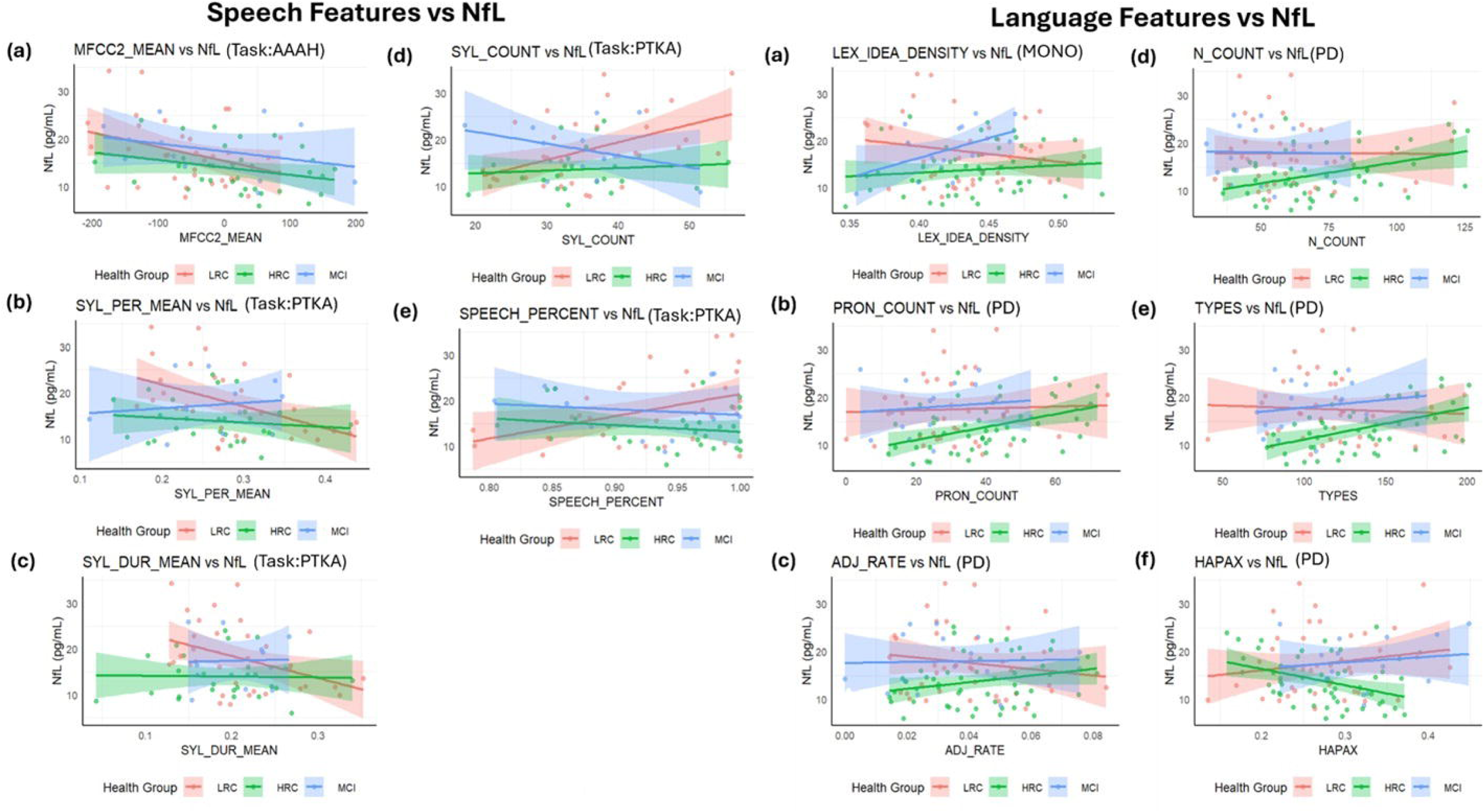
Regression plots for speech and language features with NfL. MFCC2_MEAN, mean of the second mel-frequency cepstral coefficient; SYL_COUNT, total syllable count; LEX_IDEA_DENSITY, lexical idea density; N_COUNT, total noun count; PRON_COUNT, pronoun count; TYPES, number of unique word types; ADJ_RATE, adjective production rate; HAPAX, hapax legomena; SYL_PER_MEAN, mean time between syllable onsets; SYL_DUR_MEAN, mean syllable duration; SPEECH_PERCENT, percentage of speech. AAAH, s*ustained vowel phonation; PTKA, diadochokinetic speech repetition task; MONO, monologue; PD, picture description .LRC, low-risk cognitively normal group; HRC, high-risk cognitively normal group; MCI, mild cognitive impairment. NfL, Neurofilament light chain*.

**Table 2:** Descriptive Statistics for participants.

| **Descriptive Statistics by Groups** |  |  |  |
| --- | --- | --- | --- |
| **Groups** | **LRC** | **HRC** | **MCI** |
| **N** | 49 | 60 | 18 |
| **Female** | 24 | 43 | 7 |
| **Male** | 25 | 17 | 11 |
| **Age** | 66.71 ± 6.6 | 65.5 ± 5.42 | 69.28 ± 5.23 |
| **MoCA score** | 27.23 ± 1.59 | 27.83 ± 1.8 | 23.54 ± 2.7 |
| **CDR score** | 0 | 0 | 0.5 |
| **First degree relative with AD** | Negative | Positive/<br>Suspected | - |
| **Substantial subjective memory complaints** | Negative | Positive | - |
| **Status for APOE ε4 gene allele** | Non-Carrier | Carrier (at<br>least one) | - |
| **P-tau217 (pg/ml)** | 0.33 ± 0.27 | 0.48 ± 0.31 | 0.67 ± 0.57 |
| **NfL (pg/ml)** | 17.78 ± 6.92 | 13.89 ± 4.9 | 17.69 ± 5.37 |

**Note:** LRC= Low Risk Cognitively normal, MCI= Mild Cognitive Impairment, as determined by a clinical diagnosis of MCI, RBANS Delayed Memory Index (DMI) score ≤ 85, and a positive previous biomarker score (PET imaging or CSF study). N = Number of participants. MoCA = Montreal cognitive assessment. CDR = Clinical dementia rating. APOE = Apolipoprotein E. NfL = Neurofilament light chain. Values for Age, P-tau 217 and NfL are presented as Mean ± SD.

**Table 3:**
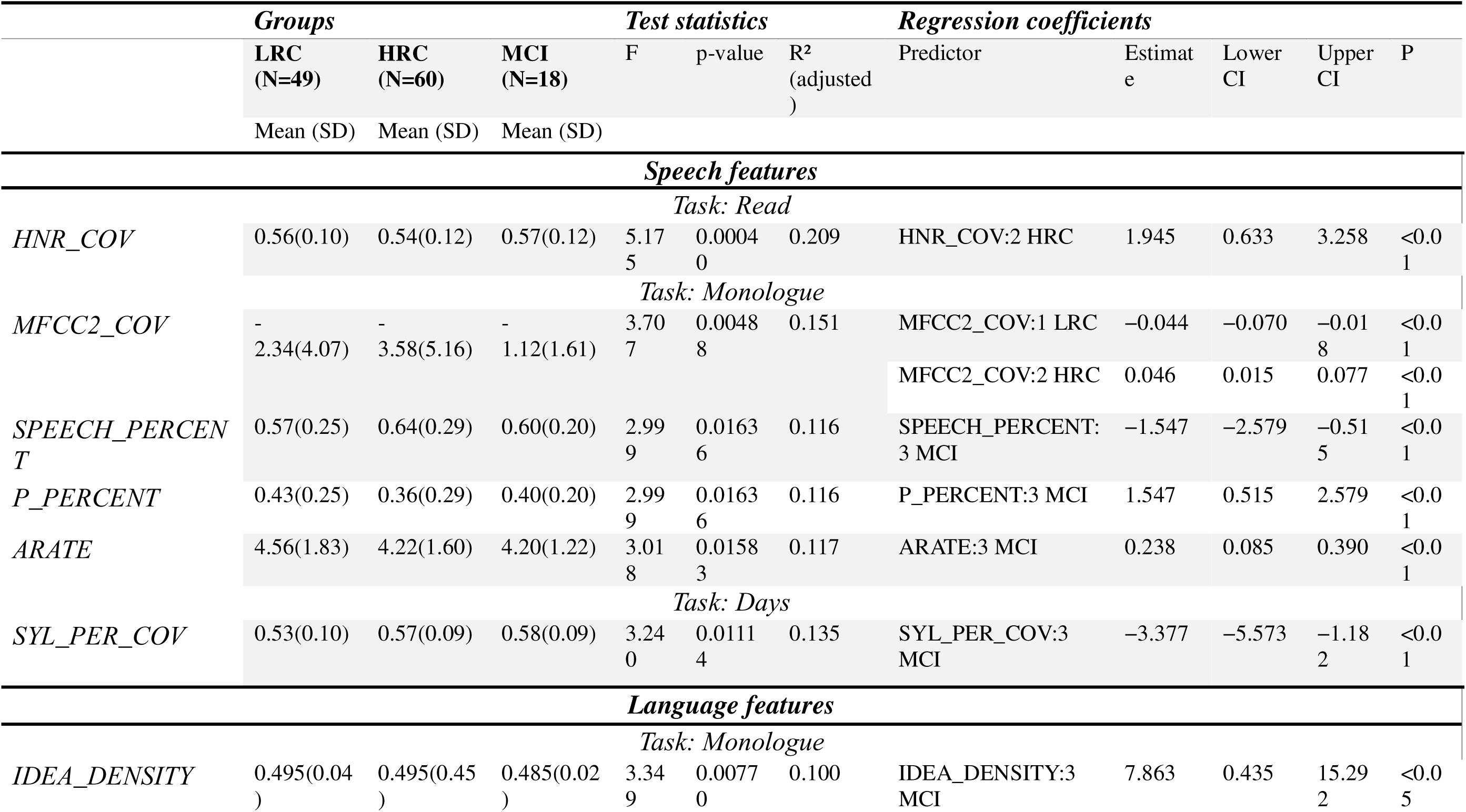
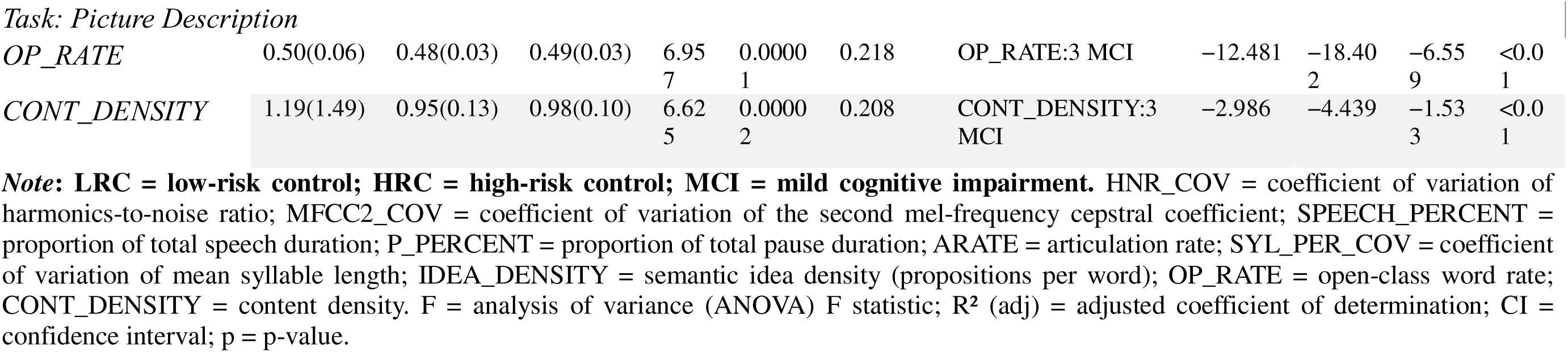
Test Statistics and regression Coefficients for speech and language features with p-tau217.

**Table 4:**
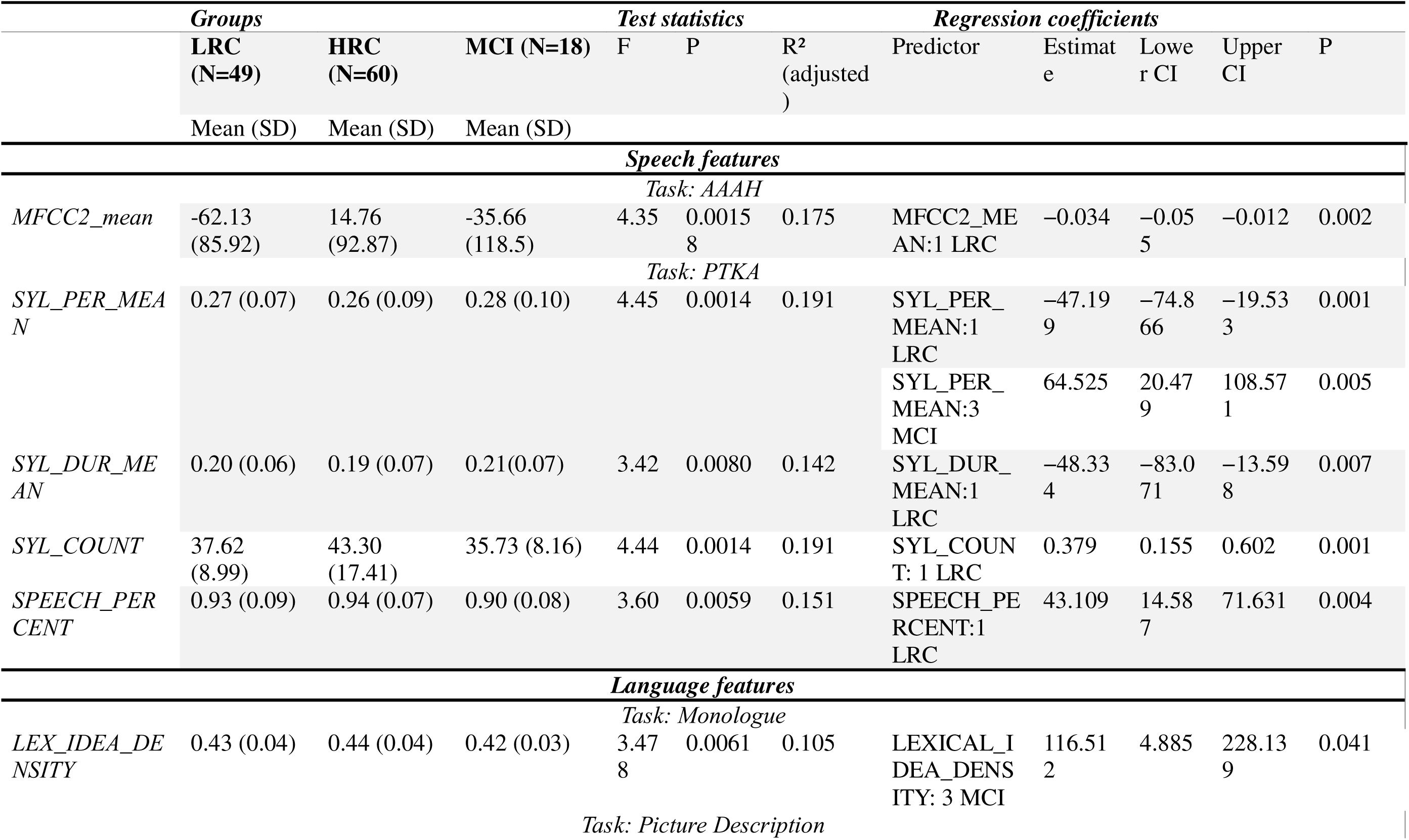
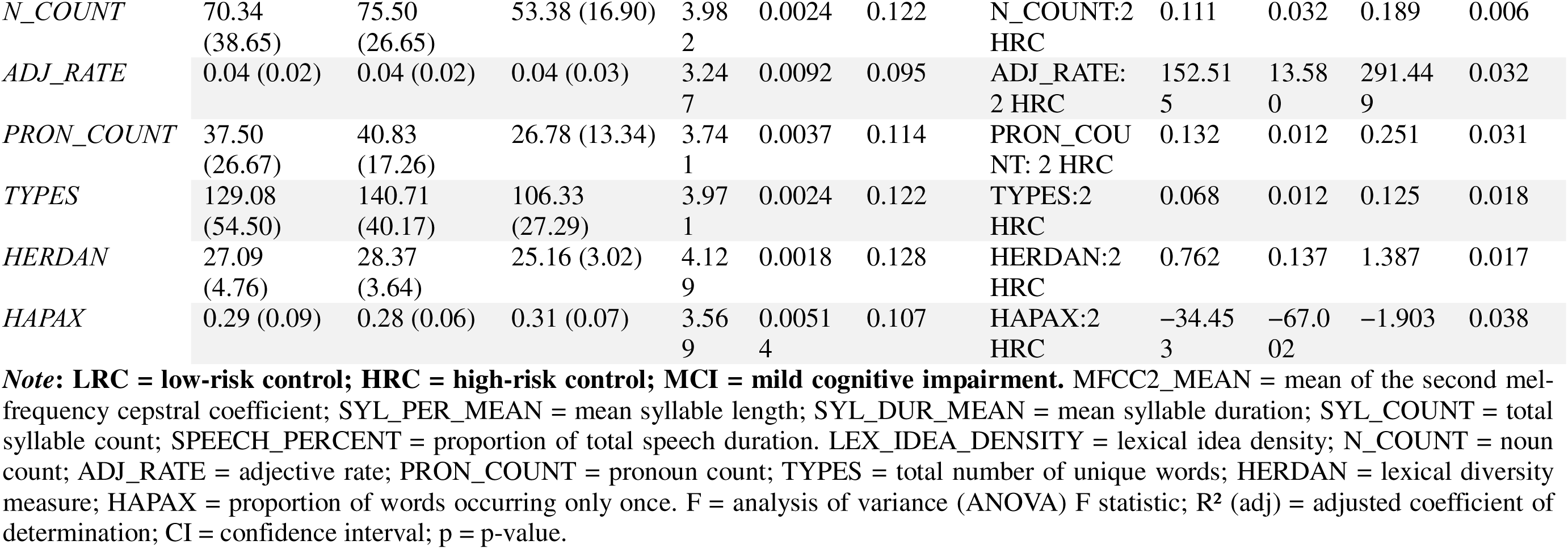
Test Statistics and regression Coefficients for speech and language features with NfL.

## 3. Results

### 3.1 Study Participants and Clinical Classification

After excluding cases with missing biomarker, speech, language, or APOE genotyping data, 127 participants were included in the analysis: 49 in the Low-Risk Cognitively Normal group (LRC), 60 in the High-Risk Cognitively Normal group (HRC), and 18 with Mild Cognitive Impairment (MCI). The mean age ranged from 65.5□±□5.4 years in HRC to 69.3□±□5.2 years in MCI. MoCA scores averaged 27.2□±□1.6 in LRC, 27.8□±□1.8 in HRC, and 23.5□±□2.7 in MCI. Mean p-tau217 levels were 0.33□±□0.27 pg/mL in LRC, 0.48□±□0.31 pg/mL in HRC, and 0.67□±□0.57 pg/mL in MCI. Mean NfL levels were 17.8□±□6.9 pg/mL in LRC, 13.9□±□4.9 pg/mL in HRC, and 17.7□±□5.4 pg/mL in MCI (Table 2).

### 3.2 Associations Between Speech and Language Features and p-tau217

Significant group differences were observed in several speech features. HNR_COV during the read task was elevated in the HRC group and positively predicted p-tau217 (F(1,6) = 5.175, p = 0.0004, adj. R² = 0.209; Estimate = 1.945, 95% CI: 0.633 to 3.258). MFCC2_COV during the monologue task was lower in the LRC group (Estimate = -0.044, 95% CI: -0.070 to -0.018) and higher in HRC (Estimate = 0.046, 95% CI: 0.015 to 0.077), both significantly predicting p-tau217 (F = 3.707, p = 0.0048, adj. R² = 0.151).

In MCI, reduced speech percent (Estimate = -1.547, 95% CI: -2.579 to -0.515) and increased pause percent (Estimate = 1.547, 95% CI: 0.515 to 2.579) were predictive of higher p-tau217 levels (F = 2.999, p = 0.016, adj. R² = 0.116). Articulation rate also showed significant group differences (F = 3.018, p = 0.0158, adj. R² = 0.117), with higher values associated with elevated p-tau217 in MCI (Estimate = 0.238, 95% CI: 0.085 to 0.390). SYL_PER_COV from the days task was lower in MCI and negatively associated with p-tau217 (F = 3.240, p = 0.011, adj. R² = 0.135; Estimate = -3.377, 95% CI: -5.573 to -1.182).

Language features also showed group differences. Idea density from the monologue task predicted p-tau217 in MCI (F = 3.349, p = 0.0077, adj. R² = 0.100; Estimate = 7.863, 95% CI: 0.435 to 15.292). From the picture description task, open-class word rate (F = 6.957, p = 0.00001, adj. R² = 0.218; Estimate = -12.481, 95% CI: -18.402 to -6.559) and content density (F = 6.625, p = 0.00002, adj. R² = 0.208; Estimate = -2.986, 95% CI: -4.439 to -1.533) were negatively associated with p-tau217 in MCI (Table 3, Figure 1).

### 3.3. Associations Between Speech and Language Features and NfL

Across groups, MFCC2_mean during sustained phonation differed significantly (F = 4.35, p = 0.0016, adj. R² = 0.175), with a negative association observed in LRC (Estimate = -0.034, 95% CI: -0.055 to -0.012, p = 0.002). Diadochokinetic task features, including SYL_PER_MEAN (F = 4.45, p = 0.0014, adj. R² = 0.191) showed opposing associations: negative in LRC (Estimate = -47.199) and positive in MCI (Estimate = 64.525, both p < 0.01). Syllable duration and count (SYL_DUR_MEAN, SYL_COUNT) were also predictive in LRC (p < 0.01), as was speech percent (Estimate = 43.109, 95% CI: 14.587 to 71.631).

Articulation rate significantly predicted NfL in the HRC group (Estimate = 4.550, 95% CI: 1.474 to 7.626, p = 0.004). Language measures including lexical idea density in MCI (Estimate = 116.512, 95% CI: 4.885 to 228.139, p = 0.041), and multiple lexical features in HRC from the picture description task like noun count, adjective rate, pronoun count, type count, Herdan’s C, and Hapax rate, all showed significant associations with NfL (Table 4, Figure 2).

## 4. Discussion

We investigated whether objective digital speech and language features are associated with blood-based biomarkers of Alzheimer’s disease pathology, p-tau217 and neurofilament light chain (NfL), in a population spanning low risk to mild cognitive impairment (MCI). Our core hypotheses were that speech and language measures would differ between risk groups, and predict biomarker levels, particularly in those with elevated AD pathology. Broadly, both hypotheses were supported: group differences and significant associations with biomarkers were observed across several acoustic and linguistic domains.

Variability in voice quality and articulation, as measured by harmonic-to-noise ratio and MFCC2, was significantly related to p-tau217 and NfL. Increased voice quality variability in the high-risk cognitively normal group may reflect subtle impairments in vocal fold control, possibly due to early tau-related changes affecting the motor speech system ^18^. MFCC2 variability was positively associated with p-tau217 in the MCI group, potentially indicating disrupted articulatory coordination or impaired prosodic modulation ^19,20^, which are early markers of cognitive decline ^21,22^. Notably, mean MFCC2 was negatively associated with NfL in the low risk control group, which could suggest that even in the absence of cognitive symptoms, increased neurodegeneration corresponds to diminished spectral richness, implicating early and possibly subclinical involvement of sensorimotor speech networks ^19,23^.

Variability in syllable timing, measured as the coefficient of variation of syllable lengths was higher in MCI but decreased as p-tau217 increased, potentially reflecting a shift from effortful but flexible speech toward more automatized, monotonous output, a hallmark of progressing AD pathology ^24,25^. Increased uniformity in syllable timing may reflect reduced cognitive flexibility and speech planning capacity ^6^.

In the linguistic domain, lower idea density, content density, and open-class word use were associated with higher p-tau217 in MCI, supporting our prediction that reduced linguistic richness accompanies tau pathology. Practically, a speaker with lower scores in these measures may produce speech that is less informative, more repetitive, or semantically sparse. For example, sentences may contain fewer unique ideas, rely more on filler words or closed-class words (e.g., ‘and’, ‘the’), and convey less meaningful content overall. These results are consistent with prior research linking lexical simplification to early semantic decline in AD ^26,27^. Declines in lexical diversity were also associated with NfL, suggesting that broader neuronal loss, not just tau accumulation, may impair language generation capacity. Interestingly, lexical idea density positively predicted NfL in MCI, which may reflect individual variability in educational or linguistic reserve that buffers against neurodegeneration ^28,29^.

While most findings aligned with our hypotheses, some discrepancies emerged. For example, speech percent was higher in LRC individuals with elevated NfL, contrary to expectations. One possible explanation is that neurodegeneration initially impacts subtle acoustic variability before affecting gross measures like speech duration. Similarly, some inconsistencies in pause metrics across groups may reflect the influence of cognitive reserve or motivational factors in performance.

Overall, the pattern of results supports a mechanistic link between speech motor control, expressive language, and the neurobiology of Alzheimer’s disease (AD). Tau pathology appears more closely associated with changes in timing and fluency, consistent with the anatomical distribution of tau aggregates in the medial temporal and association cortices ^30,31^. Whereas elevated NfL reflects diffuse axonal injury and broader disease severity, which may secondarily influence motor-speech control ^32,33^. The speech and language alterations observed likely reflect dysfunction in cortical regions critical for speech production, lexical retrieval, and discourse processing, including the superior ^34^, middle ^35^, and inferior temporal gyri ^36^, anterior temporal lobe ^37^, inferior frontal gyrus (IFG), medial temporal lobe, and inferior parietal lobule (IPL) ^38–40^. Many of these regions exhibit volume loss or disrupted connectivity in MCI and AD ^41^, overlapping with areas where tau and NfL pathology are prominent ^42,43^. Reduced MFCC2 variability corresponds to higher p-tau217, suggesting impaired articulatory coordination and prosodic modulation linked to superior temporal and motor regions ^44^. Slower articulation rate is associated with both NfL and p-tau217, reflecting compromised motor planning and executive control supported by the IFG, premotor cortex, and supplementary motor areas ^45,46^. Lower lexical diversity and density relate to higher p-tau217, implicating degeneration in anterior and middle temporal areas and IPL ^47^, whereas increased uniformity in syllable timing tracks with both biomarkers, reflecting reduced cognitive flexibility and speech planning due to frontal-parietal network disruption ^48,49^. These findings indicate that speech-derived measures potentially mirror underlying cortical and axonal pathology, highlighting their potential as sensitive, non-invasive markers of early AD progression.

Clinically, these findings yield a subset of speech and language features with potential utility for early screening and monitoring of Alzheimer’s disease risk. Acoustic measures, particularly variability in harmonic-to-noise ratio and MFCC2, were sensitive to both tau-related changes and neurodegeneration. Increased MFCC2 variability in individuals with MCI or cognitively normal but high-risk profiles may signal early tau-related dysfunction, while lower mean MFCC2 values in low-risk individuals were linked to higher NfL, possibly suggesting early subclinical axonal injury. Slower articulation rate, derived from both the syllable repetition task and monologue task, was observed in the MCI group compared to controls and predicted higher NfL levels, underscoring its role as a potential marker of ongoing neurodegeneration. Linguistic features derived from the monologue and picture description tasks, including idea density, content density, and open-class word use, were also reduced in MCI and associated with higher p-tau217, consistent with early declines in semantic store and access. Together, these results suggest it may be possible to enhance identification of individuals at risk for or in the early stages of Alzheimer’s disease pathology via acoustic markers of vocal tract dynamics and timing (e.g., lower MFCC2, slower articulation rate) obtained from sustained vowel, syllable repetition, and monologue tasks. Lexical diversity (variety of unique words produced) and lexical density (proportion of content words relative to total words) from monologue and picture description tasks showed higher scores in cognitively intact individuals with lower p-tau217 levels, suggesting that richer, more information-dense speech may help exclude significant tau burden, while lower values could signal subtle linguistic vulnerability despite preserved cognitive performance. Future research will extend these findings in longitudinal cohorts to track their sensitivity to disease progression and establish minimal thresholds of change and minimal clinically important differences (MCIDs) that can be applied in clinical practice.

### 4.1 Limitations

The cross-sectional design limits our capacity to establish the link between speech changes over time and biomarker progression. The modest sample size, particularly in the MCI group, limits generalizability. Educational background, and first language were not controlled, which may have introduced variance in language features. Acoustic features may also have been influenced by recording conditions, microphone variability, or analysis settings, which are known to affect reliability of speech measures ^50^. Differences in data modality (biochemical assays vs. computationally derived speech measures) also complicate direct interpretation of associations. Finally, while associations with biomarkers were observed, these do not yet translate into clinical thresholds or prognostic utility. Establishing longitudinal sensitivity and clinically meaningful thresholds will be the focus of our future analysis.

## 5. Conclusion

Data suggests speech and language features including spectral, prosodic, and lexical variables are associated with plasma levels of p-tau217 and NfL in individuals at risk for AD. The link between behavior and underlying pathology underscores the potential use of digital speech analysis to support early detection, stratification, and monitoring of neurodegenerative changes. As speech can be collected remotely through active and passive protocols, it holds promise as a scalable screening tool. Future work should focus on longitudinal validation, multimodal integration with imaging and cognitive measures, and deployment in real-world clinical and community settings.

## Data Availability

All data produced in the present study are available upon reasonable request to the authors

## Acknowledgments

This research was funded by the University of Melbourne Graduate Research Scholarship, awarded to support graduate students conducting advanced research at the University.

## Ethical approval and consent statement

The study adhered to the tenets of the Declaration of Helsinki, and informed consent from all subjects was obtained prior to experimental data collection after explanation of the nature and possible consequences of the study. The study was part of the Atlas of Retinal Imaging in Alzheimer’s Study (ARIAS; PJS served as principal investigator for ARIAS) which took place at the University of Rhode Island and Butler Hospital Memory and Aging Program, Providence, RI between 2020 and 2022, and was approved by the BayCare Institutional Review Board (IRB).

